# Pretreatment CTP collateral parameters predict good outcomes in successfully recanalized middle cerebral artery distal medium vessel occlusions

**DOI:** 10.1101/2023.07.10.23292483

**Authors:** Vivek Yedavalli, Omar Hamam, Meisam Hoseinyazdi, Elisabeth Breese Marsh, Raf Llinas, Victor Urrutia, Richard Leigh, Fernando Gonzalez, Risheng Xu, Justin Caplan, Judy Huang, Hanzhang Lu, Max Wintermark, Jeremy Heit, Adrien Guenego, Greg Albers, Kambiz Nael, Argye Hillis

## Abstract

**Background:** Distal medium vessel occlusions (DMVOs) account for a large percentage of vessel occlusions resulting in acute ischemic stroke (AIS) with disabling symptoms. We aim to assess whether pretreatment CT Perfusion collateral status (CS) parameters can serve as imaging biomarkers for good clinical outcomes prediction in successfully recanalized middle cerebral artery (MCA) DMVOs.

**Methods:** We performed a retrospective analysis of consecutive patients with AIS secondary to primary MCA-DMVOs who were successfully recanalized by mechanical thrombectomy (MT) defined as modified thrombolysis in cerebral infarction (mTICI) 2b, 2c, or 3. We evaluated the association between cerebral blood volume (CBV) index and hypoperfusion intensity ratio (HIR) independently with good clinical outcomes (modified Rankin score (mRS) 0-2) using Spearman rank correlation, logistic regression, and ROC analyses.

**Results:** From 8/22/2018 to 10/18/2022, 60 consecutive patients met our inclusion criteria (mean age 71.2 +– 13.9 years old [mean+-SD], 35 female). CBV index (r = –0.693, p < 0.001) and HIR (0.687, p < 0.001) strongly correlated with 90-day mRS. A CBV index >= 0.7 ((OR 2.27 [6.94 – 21.23], p = 0.001)) and absence of prior stroke (0.13 [0.33 – 0.86]), p = 0.024) were independently associated with good outcomes. ROC analysis demonstrated good performance of CBV Index in predicting good 90-day mRS (AUC 0.73, p = 0.003) with a threshold of 0.7 for optimal sensitivity (71% [52.0-85.8%]) and specificity (76% [54.9 – 90.6%]). HIR also demonstrated adequate performance in predicting good 90-day mRS (AUC 0.77, p = 0.001) with a threshold of 0.3 for optimal sensitivity (64.5% [45.4-80.8%]) and specificity (76.0% [54.9–90.6%]).

**Conclusions:** A CBV index ≥ 0.7 and HIR < 0.3 are independently associated with good clinical outcomes in our cohort of AIS caused by MCA-DMVOs that were successfully treated with MT.

## Introduction

Distal medium vessel occlusions (DMVOs) – defined as M2-M4 segments of the middle cerebral artery (MCA), anterior cerebral artery (ACA) segments and vertebrobasilar branches – are thought to represent 25-40% of acute ischemic stroke (AIS) and can result in disabling symptoms ^1^. The current standard of care treatment for AIS caused by DMVOs is IV thrombolysis but fails to successfully recanalize DMVOs in up to two thirds of patients^1^. With the recent technological advances, DMVOs are now increasingly being treated with mechanical thrombectomy (MT) despite current lack of consensus on guidelines^2^.

Robust collaterals have been shown to predict good outcomes in large vessel occlusions (LVOs), but the effect of collateral status (CS) on DMVOs is still an area of ongoing research. Although CT angiography-based CS grading can be performed, there is significant variability amongst readers, necessitating automated quantitative pretreatment CT perfusion (CTP) CS assessments^3, 4^. The cerebral blood volume (CBV) Index – defined as the mean rCBV obtained by dividing the average of all CBV values from the time-to-maximum (Tmax) > 6 s region within the ischemic hemisphere by the average of all CBV values from all tissue with Tmax ≤ 4 s^5^ – and the hypoperfusion intensity ratio (HIR) – defined as time to maximum (Tmax) greater than 10 seconds volume divided by the Tmax greater than 6 second volume^6–8^ – have both been previously validated as reliable quantitative CS parameters, particularly for middle cerebral artery (MCA) LVOs. Prior LVO studies have reported thresholds of greater than 0.8 for CBV index^9^ and approximately 0.4 for HIR^8^, where patients with greater than 0.8 or HIR less than 0.4 have good CS. However, despite being established in LVOs, no studies to our knowledge have assessed the optimal CBV Index threshold for MCA-DMVOs. Furthermore, the optimal threshold for HIR in MCA-DMVOs still remains underexplored.

Therefore, the primary aim of our study was to establish a threshold for CBV Index in patients with AIS due to primary MCA-DMVOs who were successfully recanalized by MT for clinical outcomes prediction. We hypothesize that the LVO threshold of approximately 0.8 for CBV index still applies to DMVOs. Our secondary aim is to also determine the predictive value of HIR in the same setting with comparison to CBV Index in order to assess the value of each CS parameter in the same cohort. For this aim, we hypothesize that a more restrictive threshold compared to the LVO threshold of 0.4 is optimal due to the longer transit time to the affected tissue. Specifically, we postulate that the previously suggested threshold of 0.3^10^ is optimal.

## Methods

### Population and Study Design

In this retrospective study, we identified consecutive patients from two comprehensive stroke centers within the Johns Hopkins Medical Enterprise (Johns Hopkins Hospital – East Baltimore and Bayview Medical Campus) from 8/22/2018 to 10/18/2022 in a continuously maintained database. This study was approved through the Johns Hopkins School of Medicine institutional review board (JHU-IRB00269637) and follows the STROBE checklist guidelines as an observational study^11^.

The inclusion criteria for this study were as follows: a) MT triage within 24 hours of symptom onset or last known well; b) diagnostically adequate multimodal pretreatment CT imaging including NCCT, CTA, and CTP; c) AIS due to a CT angiography (CTA) confirmed MCA-DMVO, specifically including M2-M4 segments of the MCA as defined by Saver et al^1^; and d) successful recanalization by MT defined as modified thrombolysis in cerebral infarction (mTICI) 2b or 3.

The study was conducted in accordance with the Declaration of Helsinki and the Health Insurance Portability and Accountability Act (HIPAA). Informed consent was waived by the institutional review boards given the retrospective study design.

The decisions to administer intravenous tissue plasminogen activator (IV tPA) and/or perform MT were made on an individual basis based on consensus stroke team evaluation per our institution protocols and were controlled for in our analyses.

### Data Collection

Baseline and clinical data were collected through electronic records and stroke center databases for each patient included demographics, risk factors for AIS (including diabetes mellitus, hypertension, coronary artery disease, atrial fibrillation), admission glucose, admission NIH stroke scale (admission NIHSS), Alberta Stroke Program Early CT Score (ASPECTS), site of occlusion, and laterality of occlusion, and IV thrombolysis administration. Additional collected parameters included number of passes, recanalization time, mTICI score; presence of complication such as hemorrhagic transformation (HT) as defined by the ECASS trial. Patients were subsequently grouped into good and poor CS based on the statistically determined optimal CBV threshold (please see below).

### Imaging Analysis

The ASPECTS scores were calculated on non-contrast CT (NCCT) and baseline CTAs were reviewed for presence and site of DMVO by an experienced neuroradiologist (VSY, 6 years of experience). The same neuroradiologist assessed the diagnostic adequacy of the CTPs where only those deemed diagnostic adequate were included in the study.

#### Imaging parameters

<u>NCCT</u>: NCCT is performed in a helical mode at 5 mm slice thickness with 0.75 mm; reconstructions (120 kilovoltage peak (kVp), 365 milliampere-seconds (mAs), Rotation Time 1 second, Acquisition Time 6 – 8 seconds, Collimation 128 x 0.6 mm, Pitch Value 0.55, Scan Direction Craniocaudal).

<u>CTA:</u> The CTA of the head and neck is performed with non-ionic iodinated contrast with 50-70 ml injected at 5-6 ml/second from the aortic arch through the vertex using a bolus triggered method at 3 mm slice thickness with 0.75 mm reconstructions. The CTA parameters are as follows: 90/150 kVp with an Sn filter, Quality Reference mAs 180, Rotation Time 0.25 seconds, Average Acquisition Time 3-5 seconds, Collimation 128 x 0.6 mm, Pitch Value 0.7, Scan Direction Craniocaudal.

<u>CTP:</u> CTP is then performed with injection of 50 ml non-ionic iodinated contrast with a 30 ml saline flush at 5-6 ml/second with anatomic coverage of 70-100 mm at 5 mm slice thickness. Parameters as follows: 70 kVP, 200 Effective mAs, Rotation Time 0.25 seconds, Average Acquisition Time 60 seconds, Collimation 48 x 1.2 mm, Pitch Value 0.7, 4D Range 114 mm x 1.5 seconds, Scan Direction Craniocaudal. CTP images are then post-processed using RAPID commercial software (IschemaView, Menlo Park) for generating Tmax maps, from which the HIR and CBV Index are calculated.

### Angiographic Assessment

The pre-MT DSA collateral assessment was performed by two experienced neuroradiologists (MH and VSY, 3 and 6 years of experience respectively) using the American Society of Interventional and Therapeutic Neuroradiology/Society of Interventional Radiology (ASITN/SIR) criteria^12^. Any discrepancies were assessed with a final score based on consensus evaluation. Grades 3 and 4 were categorized as good CS. Although Grade 2 is considered moderate CS, it was included in the poor CS group for dichotomized analysis.

The mTICI score was determined by the performing neurointerventionalist at the time of the procedure.

### Clinical Outcomes Assessment

Modified Rankin scores at discharge and 90 days (90 day mRS) in addition to discharge NIHSS were determined by a stroke neurologist or certified nurse practitioner.

### Outcome Measures

The primary outcomes were good clinical outcomes defined as 90-day mRS 0-2. The secondary outcomes included excellent outcomes (90-day mRS 0-1), discharge mRS, discharge NIHSS, and NIHSS shift (defined as the difference between discharge and admission NIHSS).

### Statistical analysis

The collected data were coded, tabulated, and statistically analyzed using IBM SPSS statistics (Statistical Package for Social Sciences) software version 28.0, IBM Corp., Chicago, USA, 2021. Quantitative data were tested for normality using Shapiro-Wilk test, then if normally distributed described as mean±SD (standard deviation) as well as minimum and maximum of the range. If data were not normally distributed, they were described as Median (1st−3rd Interquartiles) as well as minimum and maximum then compared using Mann Whitney test. Correlations between HIR and CBV Index as well as CBV Index with DSA CS and 90-day mRS were assessed by Spearman rank correlation. Receiver operating characteristics (ROC) curve was used to evaluate the performance of HIR and CBV Index where the optimal thresholds to predict 90-day mRS of 0-2 based on highest sensitivity and specificity were determined. Multivariate logistic regression analyses were also performed to assess the association of CBV index with 90-day mRS. Patients were then grouped based on the determined CBV optimal threshold. The level of significance taken at P value ≤ 0.050 was significant, otherwise was non-significant.

## Results

From 8/22/2018 to 10/18/2022, we identified 147 consecutive patients with AIS due to an MCA-DMVO. Of these 147 patients, 60 patients (mean age 71.2 +-13.9 years old [mean+-SD], 35 female) met the inclusion criteria and were included in this study. In total, 56 out of the 60 patients had available 90-day mRS. All patients had discharge mRS and discharge NIHSS available. Please see Table 1 for demographic information.

**Table (1):**
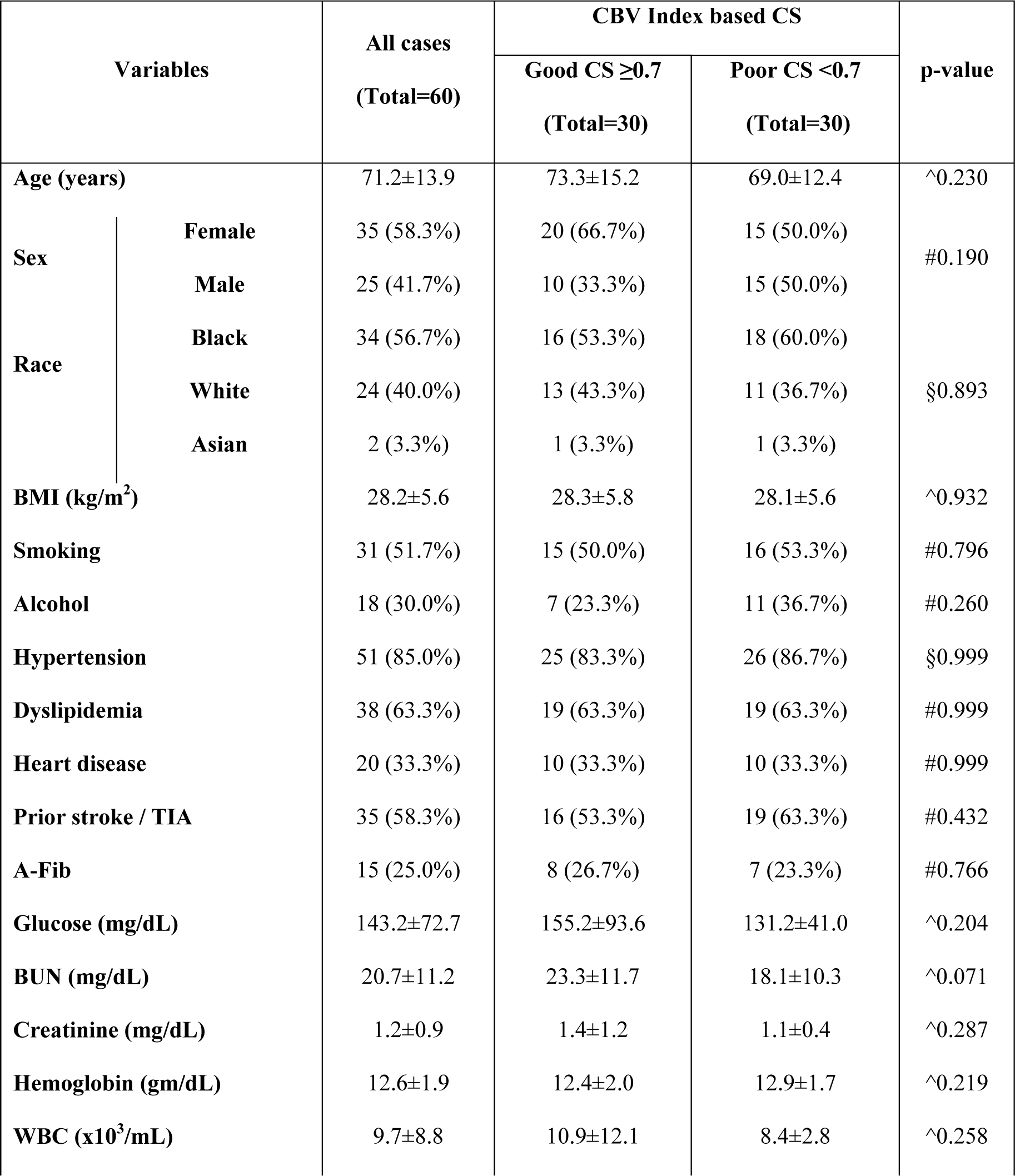
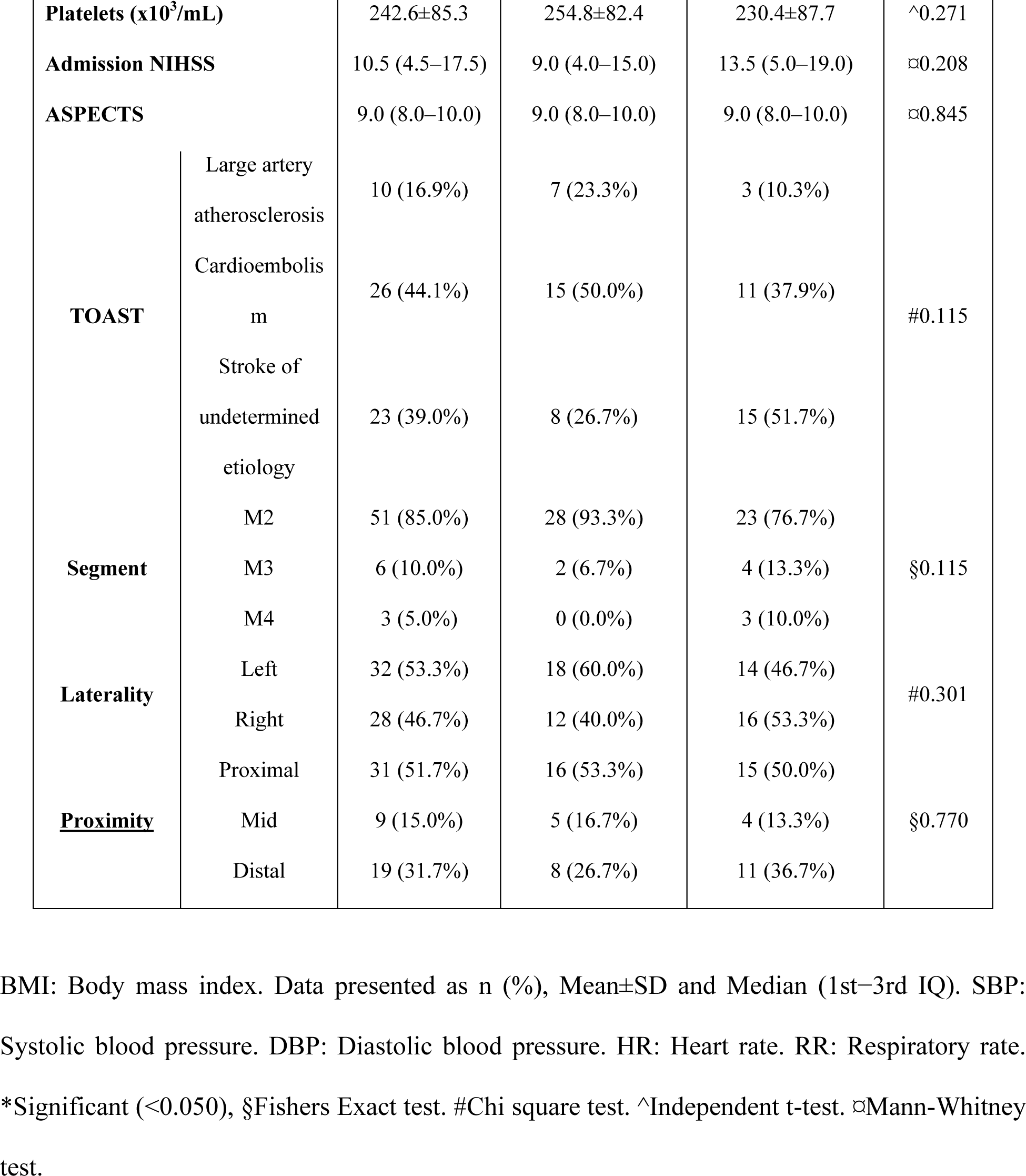
Demographic,admission findings, and vessel breakdown according to optimal CBV index threshold.

Of the 60 patients, 51 had M2 occlusions (51/60, 85%), 6 had M3s (6/60, 10%), and 3 had M4s (3/60, 5%). See Table 1.

Nineteen patients received IV thrombolysis (19/60, 31.7%) prior to MT. Forty-six (46/60, 76.7%) had pretreatment DSA that was adequate for CS assessment. Forty-three patients achieved mTICI 3 recanalization (43/60, 71.7%). Hemorrhagic transformation of any subtype was found in 15 patients (15/60, 25%). A higher percentage of moderate and good CS based on DSA were found in the CBV >= 0.7 group (4/6 vs 2/6, grade 2 and 14/20 vs 6/20, grade 3; p = 0.004) compared to the CBV < 0.7 group. Moreover, a higher percentage of poor CS based on DSA was found in the CBV < 0.7 group (4/4 vs 0.4 grade 0; 6/6 vs 0/6 grade 1; p = 0.004) versus the CBV >=0.7 group. Based on dichotomized DSA CS assessment, a higher percentage of good CS patients (19/30 vs 11/30) and a lower percentage of poor CS patients (4/16 vs 12/16) were also seen in the CBV >= 0.7 group (p = 0.013). HIR was also lower in the CBV >= 0.7 group (0.3 [median, IQR] [0.0-5.0] vs 0.5 [0.3-0.6; p = 0.001]. See Table 2.

**Table (2):**
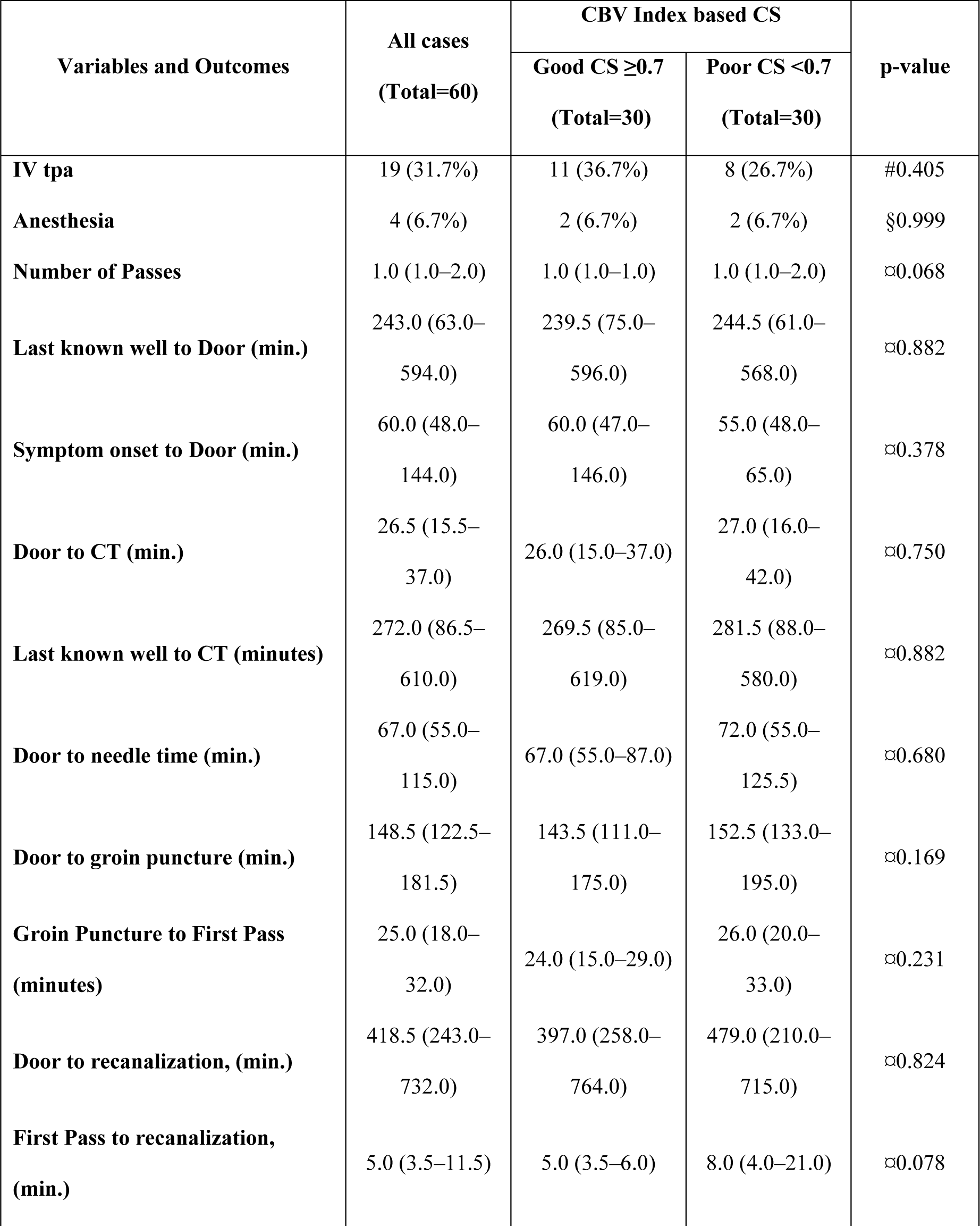
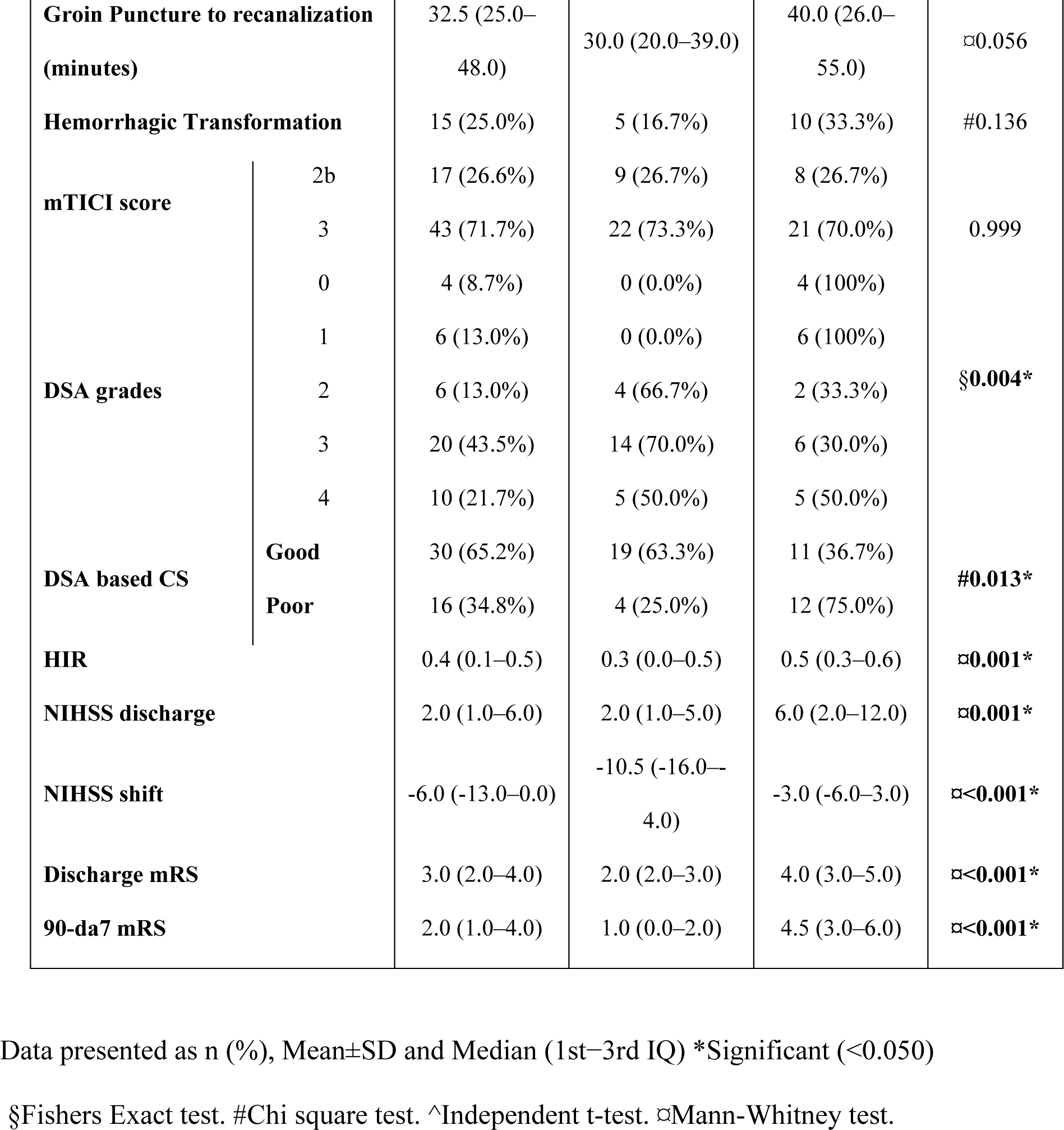
Collateral parameters, interventional parameters, and clinical outcomes based on optimal CBV index threshold.

Outcomes analysis revealed significantly lower 90 day mRS ([median, IQR] 1.0 [0.0-2.0] vs 4.5 [3.0 – 6.0], p < 0.001), discharge mRS (2.0 [2.0-3.0] vs 4.0 [3.0 – 5.0], p < 0.001), and discharge

NIHSS (2.0 [1.0-5.0] vs 6.0 [2.0 – 12.0], p = 0.001) in the CBV >= 0.7 group. A larger favorable NIHSS shift was also found in the CBV >= 0.7 group (–10.5 [–16.0– –4.0] vs –3.0 [–6.0 – 3.0], p < 0.001). Please see Table 2 for details.

### Correlation analysis

CBV Index demonstrated a strong inverse correlation with 90-day mRS (–0.693, p < 0.001) and moderate inverse correlation with HIR (–0.494, p < 0.001). HIR also demonstrated a strong positive correlation with 90-day mRS (0.687, p < 0.001). CBV Index nor HIR were significantly correlated with DSA CS assessment. Please see Table 3.

**Table (3):**
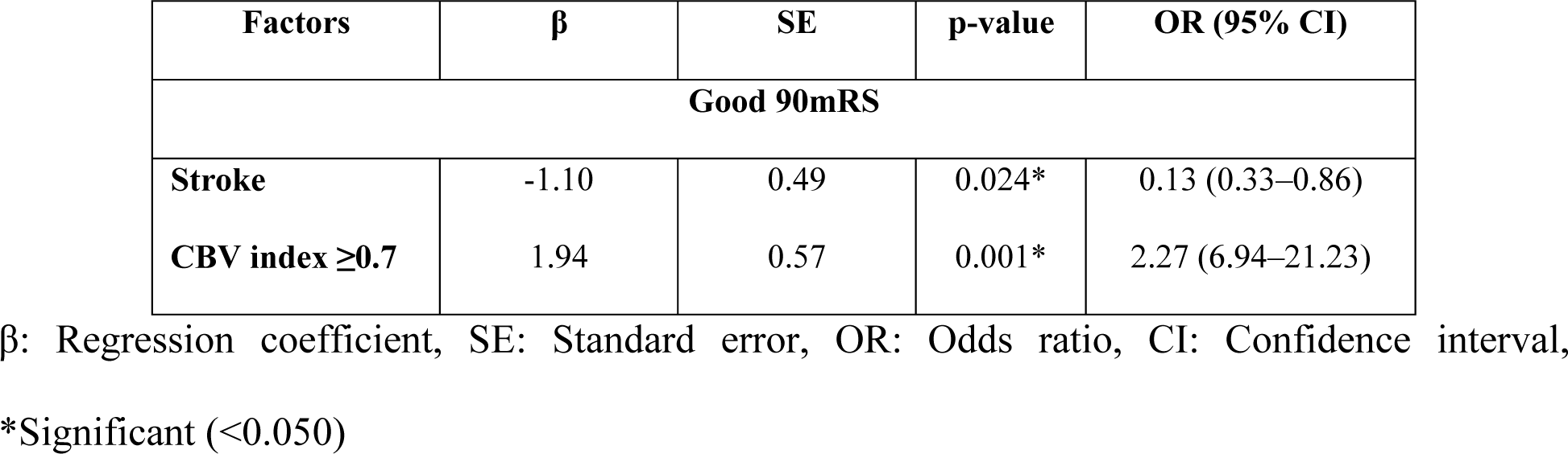
Multivariate logistic regression for predicting good clinical outcomes.

### ROC analysis

ROC analysis demonstrated adequate performance of CBV Index in predicting good 90-day mRS (AUC 0.73, p = 0.003) with a threshold of 0.7 for optimal sensitivity (71% [52.0-85.8%]) and specificity (76% [54.9 – 90.6%]). CBV index also predicted excellent 90 day mRS (AUC 0.73, p = 0.003).

HIR also demonstrated good performance in predicting good 90-day mRS (AUC 0.77, p = 0.001) with a threshold of 0.3 for optimal sensitivity (64.5% [45.4-80.8%]) and specificity (76.0% [54.9–90.6%]). HIR also predicted excellent 90-day mRS (AUC 0.741, p = 0.002). Please see Figure 1.

**Figure (1):**
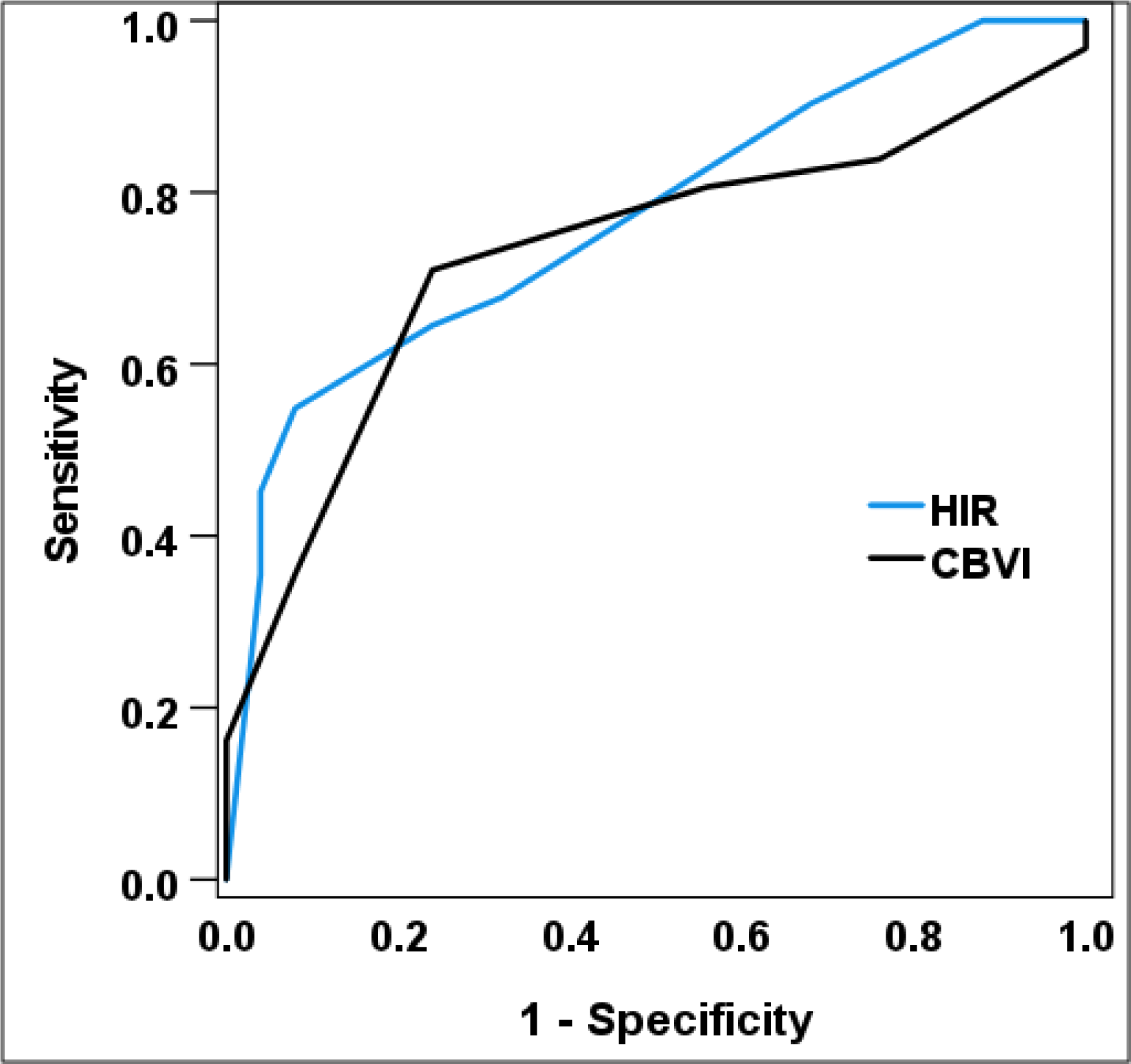
Receiver Operating Characteristics (ROC) curve for HIR and CBV in predicting good 90mRS.

No significant differences were noted between the diagnostic performances of the CBV index and HIR thresholds with respect to predicting good 90-day mRS (p = 0.558). Please see Table 4.

**Table (4):**
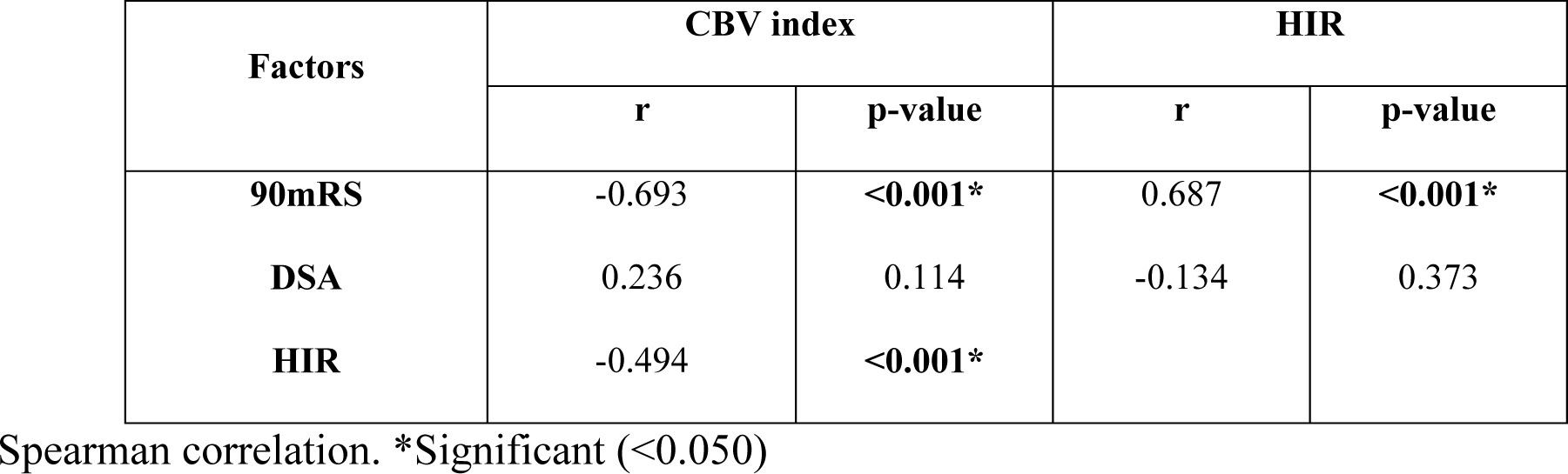
CBV index and HIR correlations with other parameters.

**Table (5):**
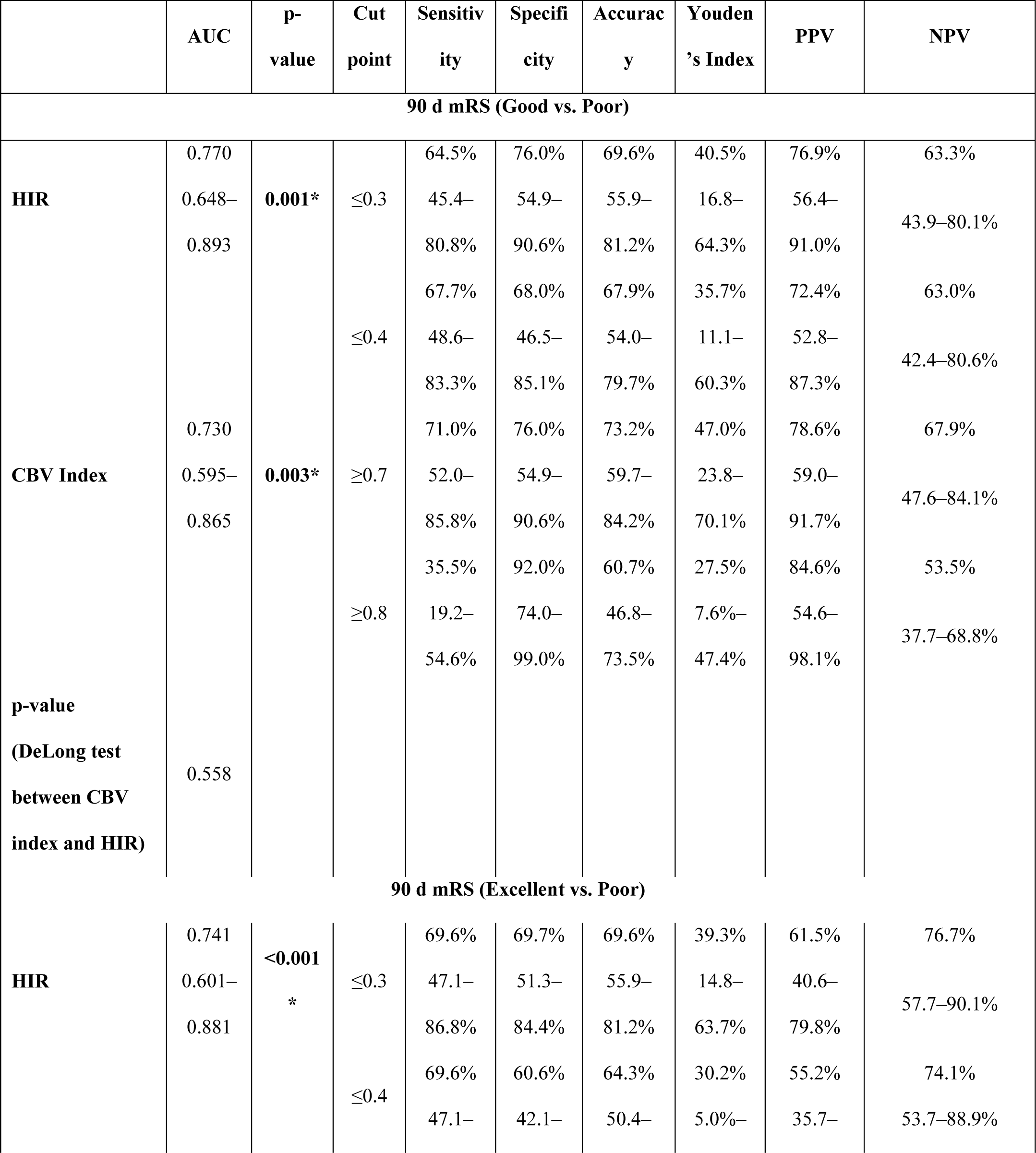
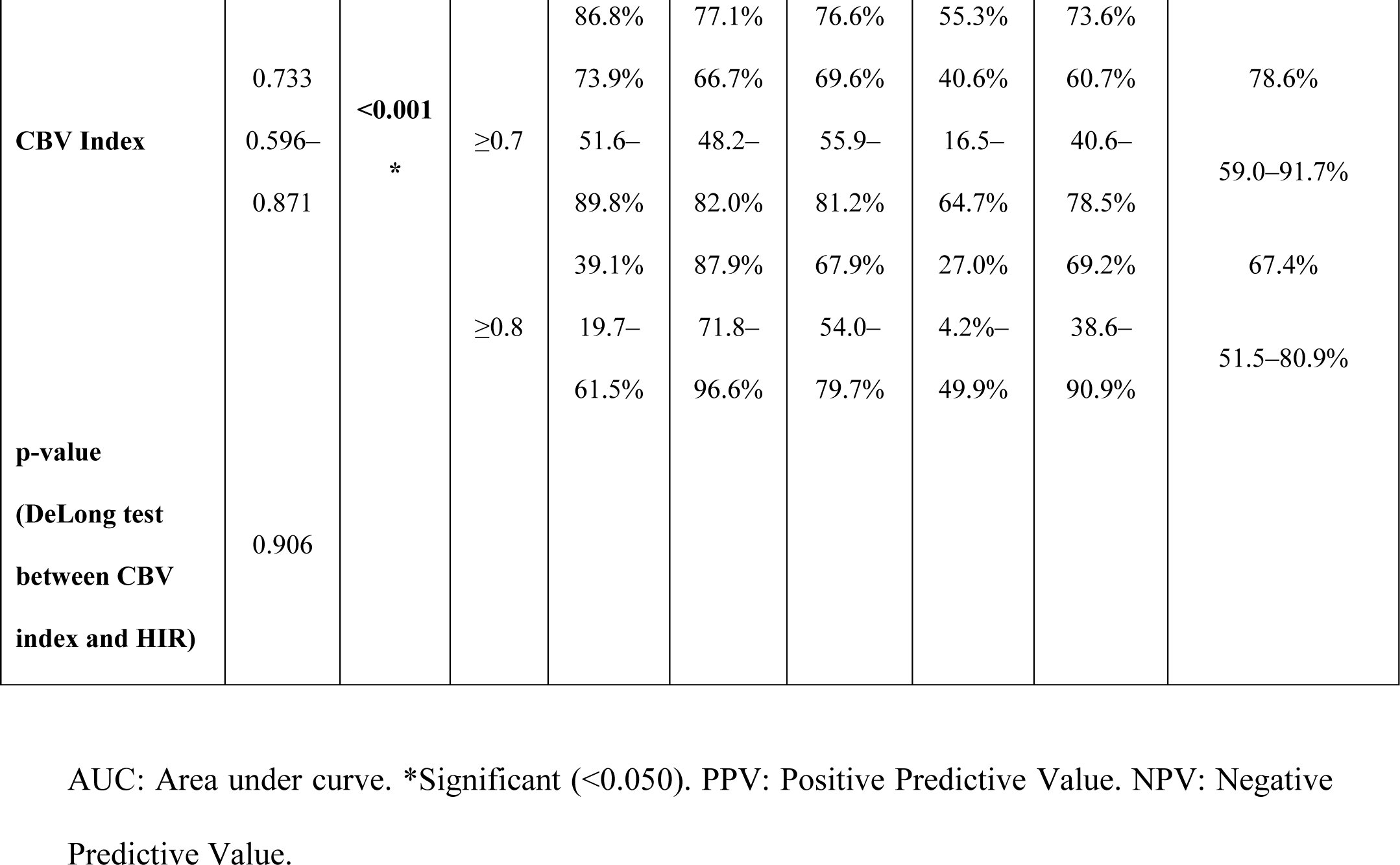
Diagnostic performance of CBV index and HIR in predicting good and excellent clinical outcomes.

### Multivariate regression analysis

A CBV index of >=0.7 (OR 2.27 [6.94 – 21.23], p = 0.001) was significantly associated with good outcomes. Furthermore, prior stroke (0.13 [0.33 – 0.86]), p = 0.024) was inversely associated with good outcomes.

## Discussion

In this study, we identify CBV index and HIR thresholds that predict good outcomes after thrombectomy treatment of MCA-DMVOs. We also demonstrate that a CBV index >= 0.7 is an independent predictor of good outcomes in this group of patients. This is the first study to our knowledge to determine a CBV index threshold for successfully recanalized AIS patients with DMVOs.

Although the current standard of care for DMVOs is IV thrombolysis, actual practice is increasingly using MT since IV thrombolysis fails to achieve recanalization in up to half of patients^13^. The advancements in endovascular technology have allowed for better navigation of these smaller caliber and often tortuous vessels. Several studies have demonstrated feasibility and safety of attempting MT in DMVOs^1, 2, 13, 14^, leading to an increased number of these patients being treated with MT.

In light of these advances in intervention, the effect of CS in DMVOs has become more relevant yet requires further investigation. As an established biomarker of infarct growth and outcomes in LVOs^7^, it stands to reason that CS similarly influences outcomes in DMVOs as well. Pretreatment CTP quantitative assessments with CBV index^5, 9, 15^ and HIR^6–8^ have been established as imaging biomarkers of CS in LVOs. However, due to the smaller volume of affected tissue and the longer transit to reach these regions, the same LVO thresholds may not apply to DMVOs, which is the main purpose of our investigation.

As an indicator of the relative blood volume within critically hypoperfused tissue, CBV index is thought to represent an indirect compensatory response to the acute occlusion through collateralization. Our results indicate that a threshold of 0.7 where a pretreatment CBV index of 0.7 or greater predicts good clinical outcomes. This threshold is lower than the established LVO threshold of greater than 0.8 for 24-hour infarct volume prediction^9^. In direct comparison with respect to DMVOs, the 0.7 threshold is more accurate (73.2% versus 60.7%) and more sensitive (71.0% versus 35.5%) than 0.8, although less specific (76% versus 92%) in predicting good clinical outcomes. We postulate that the lower CBV index for MCA-DMVOs determined in our study is due to the smaller area of affected tissue where, where compared to LVOs, a less robust compensatory response may suffice to maintain tissue viability.

Interestingly, we did not find a correlation between DSA and both CBV index and HIR, respectively. However, we found a strong inverse correlation between CBV index and 90-day mRS in addition to a strong direct correlation between HIR and 90-day mRS. This discordance between both CS parameters’ correlations with 90-day mRS and DSA may be due to a smaller sample of patients with DSA CS assessments. In our cohort, 46 patients (46/60, 76.7%) had DSAs that were imaged long enough to perform adequate CS assessments. Furthermore, despite DSA being considered the reference standard for CS assessment, prior studies are mixed on the robustness of DSA CS in predicting functional outcomes ^3, 16, 17^. It is possible that both CS parameters capture a compensatory component of CS that may translate completely to DSA but is reflected with subsequent clinical outcomes.

We also assessed the value of HIR as a predictive outcomes measure in successfully recanalized DMVOs. HIR is a well-established CS imaging biomarker in LVOs where a 0.4 threshold has been optimal^6–8^. LVOs patients with an HIR of less than 0.4, thought to represent good CS, has been correlated with good DSA collaterals in M1 occlusions^6^, validated for transferring patients for MT^8^, and predictive of infarct growth rate and clinical outcomes^7^ as well as post MT HT^18^. However, the role HIR plays in DMVOs is still being investigated.

Guenego et al most recently concluded that an HIR of less than 0.3 was associated with good CS and predicted less infarct growth in successfully recanalized DMVO cohort of 40 patients^10^. They also found that patients with an HIR equal to or greater than 0.3 had unfavorable outcomes on univariate analysis. However, this did not persist on multivariate analysis^10^. Our results are concordant with Guenego et al where we further validate the threshold of 0.3. In our cohort, patients with an HIR less than 0.3 predicted good and excellent outcomes, even doing so slightly superior to CBV index, although the difference was not significant (p = 0.558). Our study also has some notable differences compared to Guenego et al. First, our study has a larger sample size of 60 patients. Our analysis also focused on directly predicting clinical outcomes as opposed to infarct growth as a clinical outcome surrogate. In comparison to the LVO threshold of 0.4, the 0.3 threshold was slightly more accurate (69.6% versus 67.9%) and more specific (76.0% versus 68.0%), although less sensitive (64.5% versus 67.7%) in predicting good clinical outcomes. We hypothesize that this lower threshold for HIR compared to the LVO threshold of 0.4 is likely due to longer transit time to reach the affected region, necessitating a more restrictive threshold.

In addition to our CS parameter assessment, we also report that a history of prior stroke decreases the likelihood of good outcomes in these patients. Prior stroke as a predictive biomarker is underexplored within this patient population. In a study assessing medium vessel occlusions with discrepant infarct patterns, Ospel et al found a history of prior stroke in 16.4% (43/262) of patients in their cohort^19^. Our cohort had a substantially higher percentage of patients with prior stroke (58.3%, 35/60). The difference in sample size may be the reason for this discrepancy with Ospel et al. Nevertheless, prior stroke is a well-established risk factor for stroke recurrence of all types ^20–22^ and, for that reason, it is understandable that a similar trend may also apply to the MCA-DMVO population. This may be an area of future research with larger studies.

Our study has several limitations. Firstly, it is inherently limited by its retrospective design. Secondly, we focused on only MCA-DMVOs with a predominance of M2 occlusions, which may introduce a bias. The M2 occlusion predominance is most likely due to the relative proximity of the vessel, making these occlusions more amenable to MT. Thirdly, our analysis is restricted to use of one commercial software platform, which may limit generalizability.

Nevertheless, our study is strengthened by an adequate sample size of 60, given the stringent inclusion criteria of successfully recanalized MCA-DMVOs with MT. Our cohort consists of two comprehensive stroke centers serving different demographics, therefore improving generalizability.

In conclusion, the use of automated pretreatment CTP CS measures may have promise in everyday clinical practice. Currently, to our knowledge, no threshold for CBV index in the setting of DMVOs exists. Moreover, the utility of HIR in DMVOs still requires additional exploration. Our study demonstrates that, in comparison to LVO thresholds, a lower CBV index and a more restrictive HIR are associated with improved clinical outcomes in successfully recanalized MCA-DMVO patients. Given the prevalence of DMVOs, these thresholds have potential utility in everyday clinical practice as additional predictive imaging biomarkers in this group. Nevertheless, larger scale studies must be performed to further assess the strength of our results.

## Data Availability

Data can be made available upon reasonable request.

## Non-standard Abbreviations and Acronyms

Distal medium vessel occlusions = DMVOs, acute ischemic stroke = AIS, CT Perfusion = CTP, CT angiography = CTA, non-contrast CT = NCCT, collateral status = CS, middle cerebral artery = MCA, mechanical thrombectomy = MT, modified thrombolysis in cerebral infarction = mTICI, cerebral blood volume = CBV, hypoperfusion intensity ratio = HIR, modified rankin score = mRS, anterior cerebral artery = ACA, large vessel occlusion = LVO, time-to-maximum = Tmax, NIH stroke scale (NIHSS), hemorrhagic transformation = HT, intravenous tissue plasminogen activator = IV tPA, Alberta Stroke Program Early CT Score = ASPECTS, kilovoltage peak = kVp, milliampere-seconds = mAs, body mass index = BMI, systolic blood pressure = SBP, diastolic blood pressure = DBP, heart rate = HR, respiratory rate = RR.

## Acknowledgements

None

